# Association between consumption of vegetables and COVID-19 mortality at a country level in Europe

**DOI:** 10.1101/2020.07.17.20155846

**Authors:** Susana C Fonseca, Ioar Rivas, Dora Romaguera, Marcos Quijal-Zamorano, Wienczyslawa Czarlewski, Alain Vidal, Joao A Fonseca, Joan Ballester, Josep M Anto, Xavier Basagana, Luis M Cunha, Jean Bousquet

## Abstract

**Background:** Many foods have an antioxidant activity, and nutrition may mitigate COVID-19. To test the potential role of vegetables in COVID-19 mortality in Europe, we performed an ecological study.

**Methods:** The European Food Safety Authority (EFSA) Comprehensive European Food Consumption Database was used to study the country consumption of Brassica vegetables (broccoli, cauliflower, head cabbage (white, red and savoy cabbage), leafy brassica) and to compare them with spinach, cucumber, courgette, lettuce and tomato. The COVID-19 mortality per number of inhabitants was obtained from the Johns Hopkins Coronavirus Resource Center. EuroStat data were used for potential confounders at the country level including Gross Domestic Product (GDP) (2019), population density (2018), percentage of people over 64 years (2019), unemployment rate (2019) and percentage of obesity (2014, to avoid missing values). Mortality counts were analyzed with quasi-Poisson regression models to model the death rate while accounting for over-dispersion.

**Results:** Of all the variables considered, including confounders, only head cabbage and cucumber reached statistical significance with the COVID-19 death rate per country. For each g/day increase in the average national consumption of some of the vegetables (head cabbage and cucumber), the mortality risk for COVID-19 decreased by a factor of 11, down to 13.6 %. Lettuce consumption increased COVID-19 mortality. The adjustment did not change the point estimate and the results were still significant.

**Discussion:** The negative ecological association between COVID-19 mortality and the consumption of cabbage and cucumber supports the *a priori* hypothesis previously reported. The hypothesis needs to be tested in individual studies performed in countries where the consumption of vegetables is common.

## Introduction

One of the striking problems raised by the COVID-19 pandemic is the highly variable death rate between and within countries ^1^. The COVID-19 epidemic is multifactorial but, among potentially relevant factors, diet has received little attention. Many foods have an antioxidant activity ^2-4^ and it has been proposed that the role of nutrition may mitigate COVID-19 ^5^. Some countries with low COVID-19 mortality rates appear to be those with a relatively high consumption of traditional foods, many of which are fermented ^1^. It was proposed that this was associated with the antioxidant activity of the foods acting on insulin resistance ^1^.

The European Food Safety Authority (EFSA) Comprehensive European Food Consumption Database was used to study the country consumption of fermented vegetables, pickled/marinated vegetables, fermented milk, yoghurt and fermented sour milk. The negative ecological association between COVID-19 mortality and the consumption of fermented vegetables ^6^ supported the *a priori* hypothesis previously reported ^1^. In this hypothesis, we proposed that vegetables such as Brassica - with an antioxidant activity reducing insulin resistance - may also be associated with low COVID-19 mortality in countries.

To test the potential role of Brassica and some other antioxidant vegetables in the COVID-19 mortality in Europe, we performed an ecological study with Brassica vegetables (broccoli, cauliflower, head cabbage (white, red and savoy cabbage), leafy brassica) and compared them with spinach, cucumber, courgette, lettuce and tomato.

## Methods

### Selection of the database

The European Union provided the European Food Safety Authority (EFSA) Comprehensive European Food Consumption Database in 2011 as well as an update in 2015 as previously proposed.

### Selection of foods

The hypothesis raised recently proposed that diet with Brassica vegetables may explain some of the differences in the COVID-19 death rates between countries. We then selected broccoli, cauliflower, head cabbage (white, red and savoy cabbage), leafy brassica, spinach, cucumber, courgette, lettuce, and tomato as a control.

Data from the national surveys of different EU countries have been extracted from the EFSA Comprehensive European Food Consumption Database for different vegetables (Tables 1 and 2).

**Table 1:**
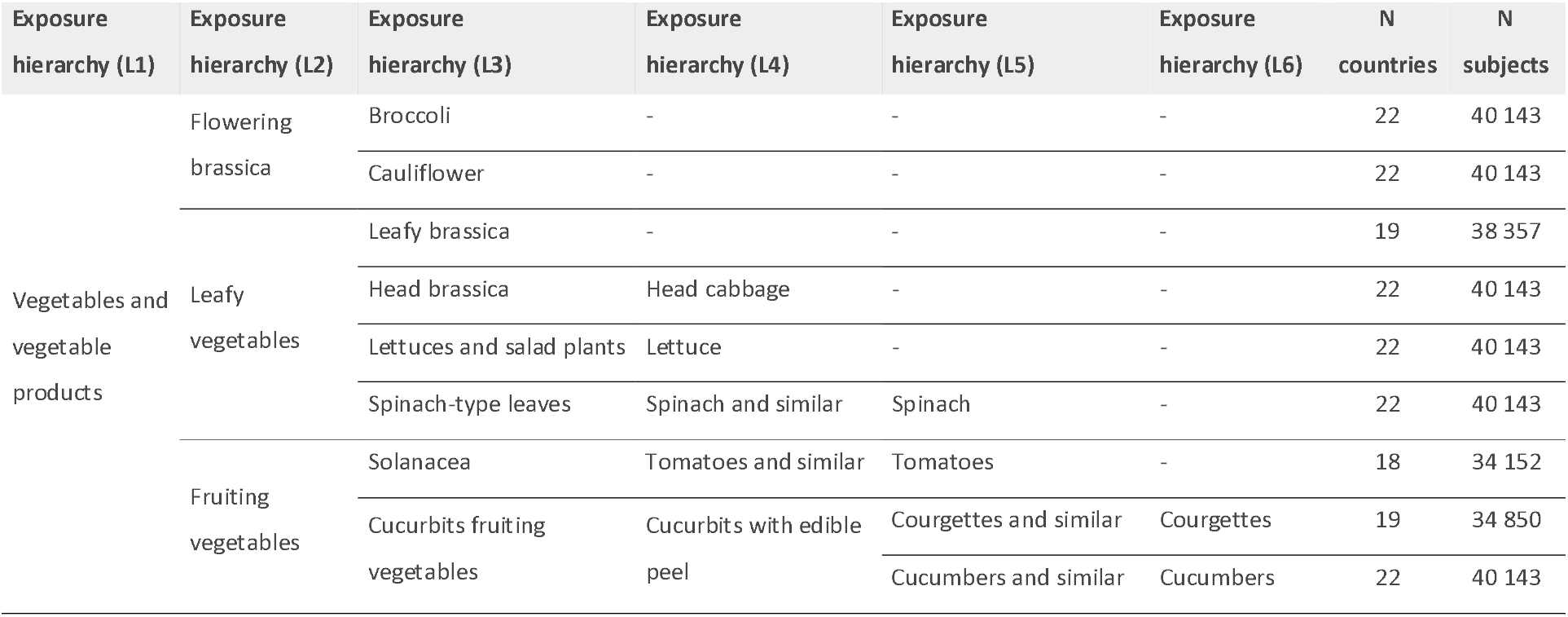
Foods tested (EFSA database)

**Table 2:** **Origin (survey and year) of consumption data obtained from the EFSA Comprehensive European Food Consumption database**

| Country | Year | Survey name | Broccoli | Cauliflower | Leafy brassica | Head cabbage | Lettuce | Spinach | Tomatoes | Courgettes | Cucumbers |
| --- | --- | --- | --- | --- | --- | --- | --- | --- | --- | --- | --- |
| Austria | 2014 | AT-NATIONAL-2016 | + | + | + | + | + | + | + | + | + |
| Belgium | 2014 | NATIONAL-FCS-2014 | + | + | + | + | + | + | + | + | + |
| Croatia | 2011 | NIPNOP-HAH-2011-2012 | + | + | + | + | + | + |  | + | + |
| Cyprus | 2014 | CY 2014-2017-LOT2 | + | + |  | + | + | + | + | + | + |
| Czech Republic | 2003 | SISP04 | + | + | + | + | + | + | + | + | + |
| Denmark | 2005 | DANSDA 2005-08 | + | + | + | + | + | + | + |  | + |
| Estonia | 2013 | DIET-2014-EST-A | + | + | + | + | + | + | + |  | + |
| Finland | 2012 | FINDIET2012 | + | + | + | + | + | + |  | + | + |
| France | 2014 | INCA3 | + | + | + | + | + | + | + | + | + |
| Germany | 2007 | NATIONAL NUTRITION SURVEY II | + | + | + | + | + | + | + | + | + |
| Greece | 2014 | GR-EFSA-LOT2 2014-2015 | + | + |  | + | + | + | + | + | + |
| Hungary | 2003 | NATIONAL REPR SURV | + | + | + | + | + | + | + | + | + |
| Ireland | 2008 | NANS 2012 | + | + | + | + | + | + | + | + | + |
| Italy | 2005 | INRAN SCAI 2005-06 | + | + | + | + | + | + | + | + | + |
| Latvia | 2012 | LATVIA_2014 | + | + | + | + | + | + | + | + | + |
| Netherlands | 2012 | FCS2016_CORE | + | + | + | + | + | + | + | + | + |
| Portugal | 2015 | IAN.AF 2015-2016 | + | + | + | + | + | + | + | + | + |
| Romania | 2012 | DIETA PILOT ADULTS | + | + |  | + | + | + | + | + | + |
| Slovenia | 2017 | SI.MENU-2018 | + | + | + | + | + | + | + | + | + |
| Spain | 2013 | ENALIA2 | + | + | + | + | + | + | + | + | + |
| Sweden | 2010 | RIKSMATEN 2010 | + | + | + | + | + | + |  |  | + |
| United Kingdom | 2008 | NDNS ROLLING PROGRAMME YEARS 1-3 | + | + | + | + | + | + |  | + | + |

### Assessment of COVID-19 mortality

We used mortality per number of inhabitants to assess the death rates as proposed by the European Center for Disease Prevention and Control (ecdc, https://www.ecdc.europa.eu/en). Raw data were used for the Poisson analysis.

Data on COVID-19 mortality were downloaded from the Johns Hopkins Coronavirus Resource Center (https://coronavirus.jhu.edu) on June 22^nd^, 2020. In addition, we downloaded data from EuroStat on: Gross Domestic Product (GDP) of the country (2019), population density (2018), percentage of people older than 64 years (2019), unemployment rate (2019) and percentage of obesity (2014, to avoid missing values).

### Statistical analysis

Mortality counts were analyzed with quasi-Poisson regression models - with log of population as an offset - to model the death rate while accounting for over-dispersion. Models were fitted separately for each of the food consumption groups. Due to the small number of countries included in the analyses, we adjusted our models using the first principal component of a principal component analysis (PCA) of the variables GDP, population density, percentage of people older than 64 years, unemployment and obesity ^7^. Adjustment was repeated using the variable most associated with mortality among the confounders, instead of using the principal component.

## Results

### Food intake in European countries

The intake of selected foods was given in g/day (Figure 2). For some of the foods, over 20 g/day are consumed in certain countries (head cabbage, lettuce, tomato, courgette and cucumber) whereas, for the others, less than 10 g/day are consumed (leafy Brassica, cauliflower, broccoli and spinach) (Figure 2).

### Association between COVID-19 mortality and food intake at the country level

Of all the variables considered, including confounders, only head cabbage and cucumber reached statistical significance in their association with the COVID-19 death rate per country, regardless of the adjustment. (Table 3 and Figures 1 and 2).

**Table 3:**
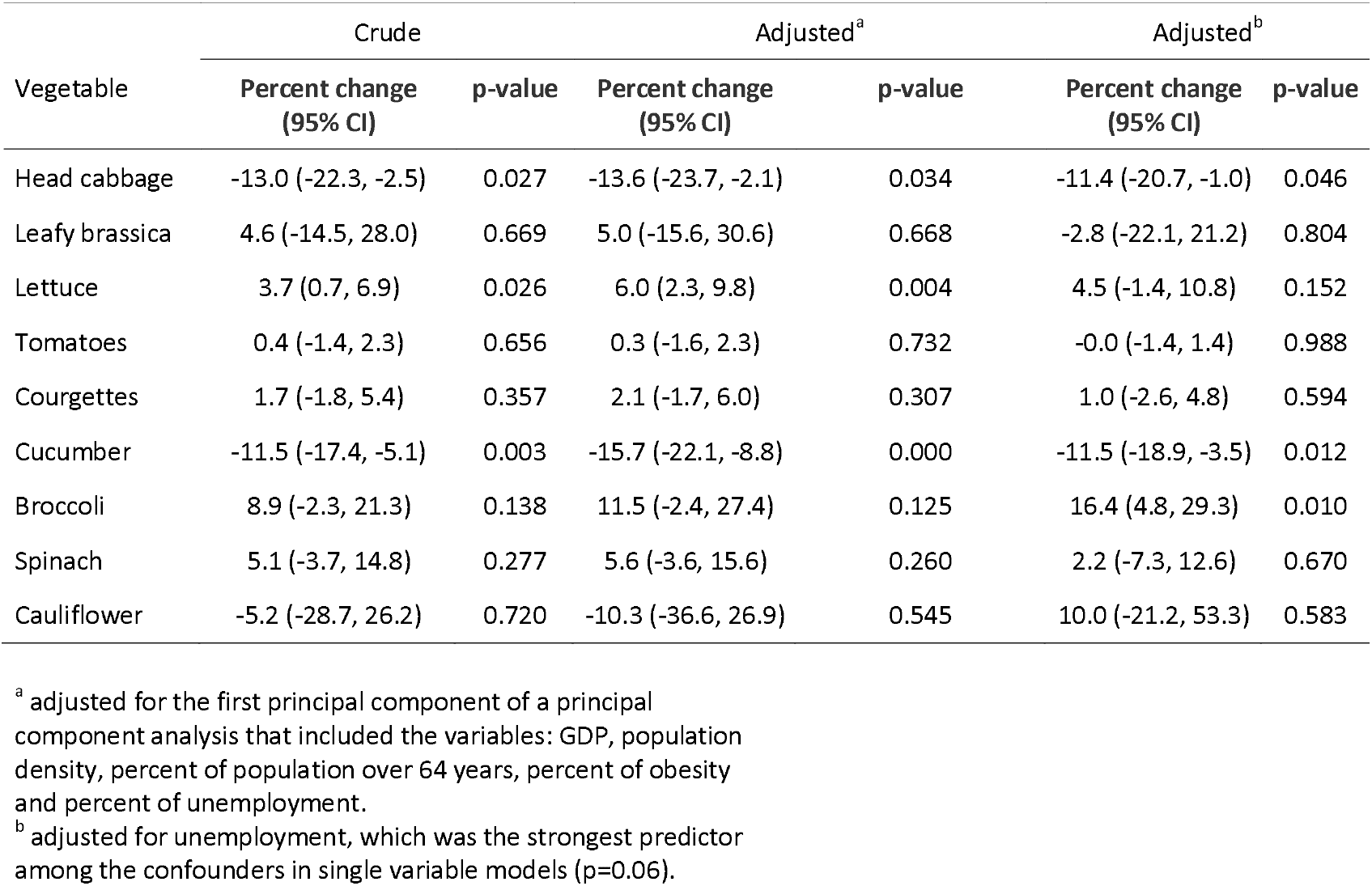
**Percent change in COVID-19 death risk (with 95% confidence intervals (CI)) as a function of diet variables. Estimations for each diet variable come from a different model**.

**Figure 1:**
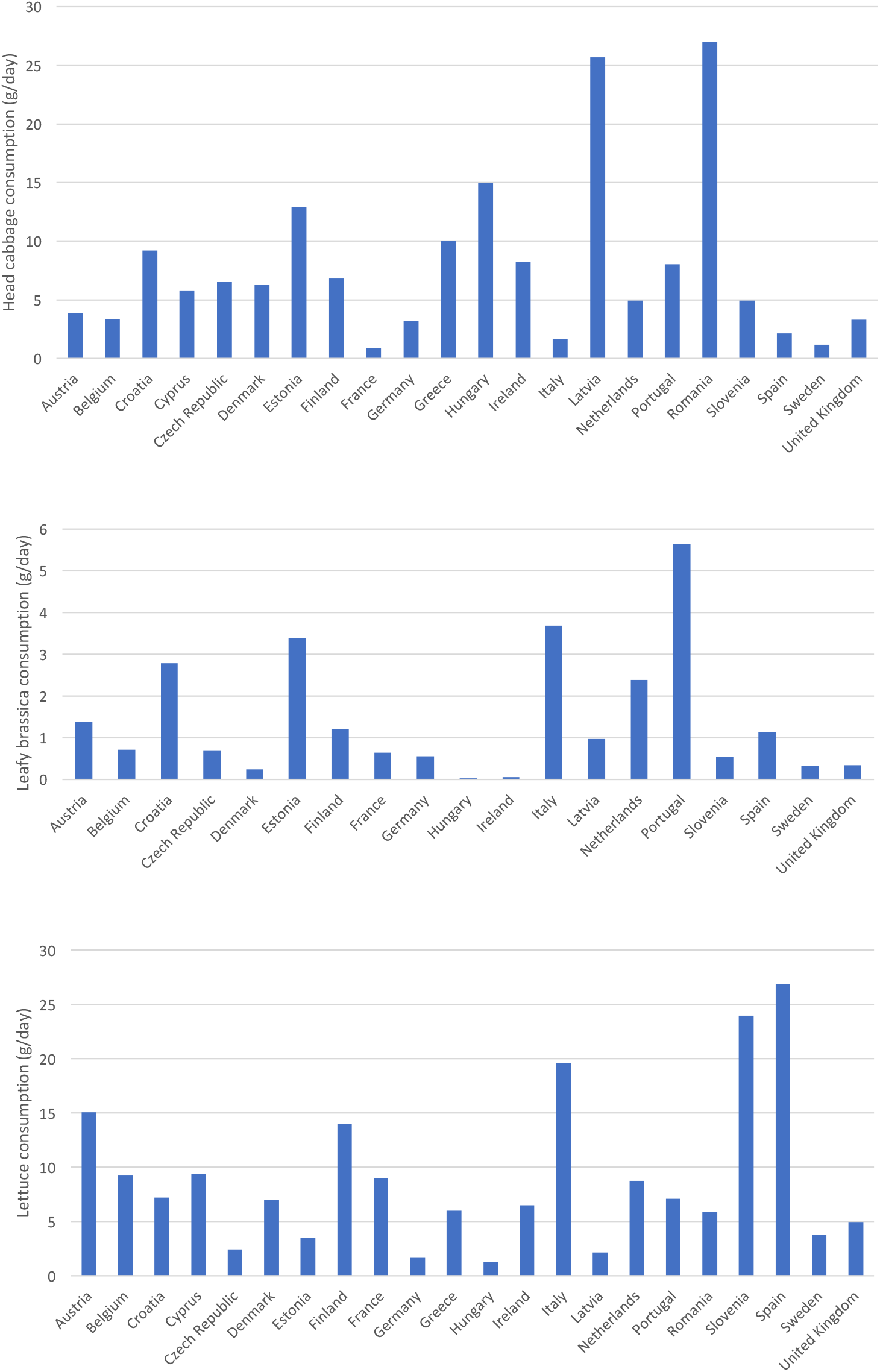
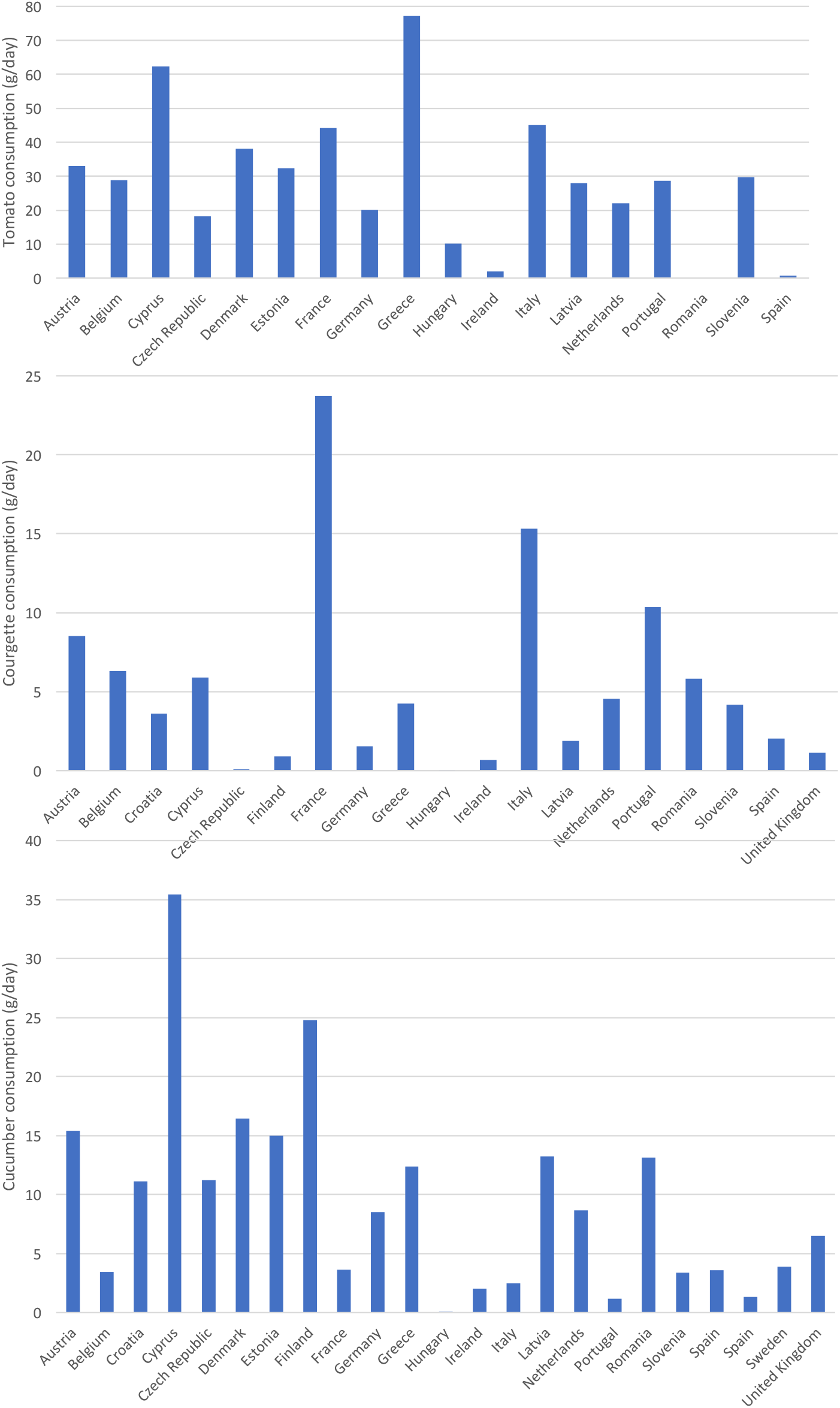
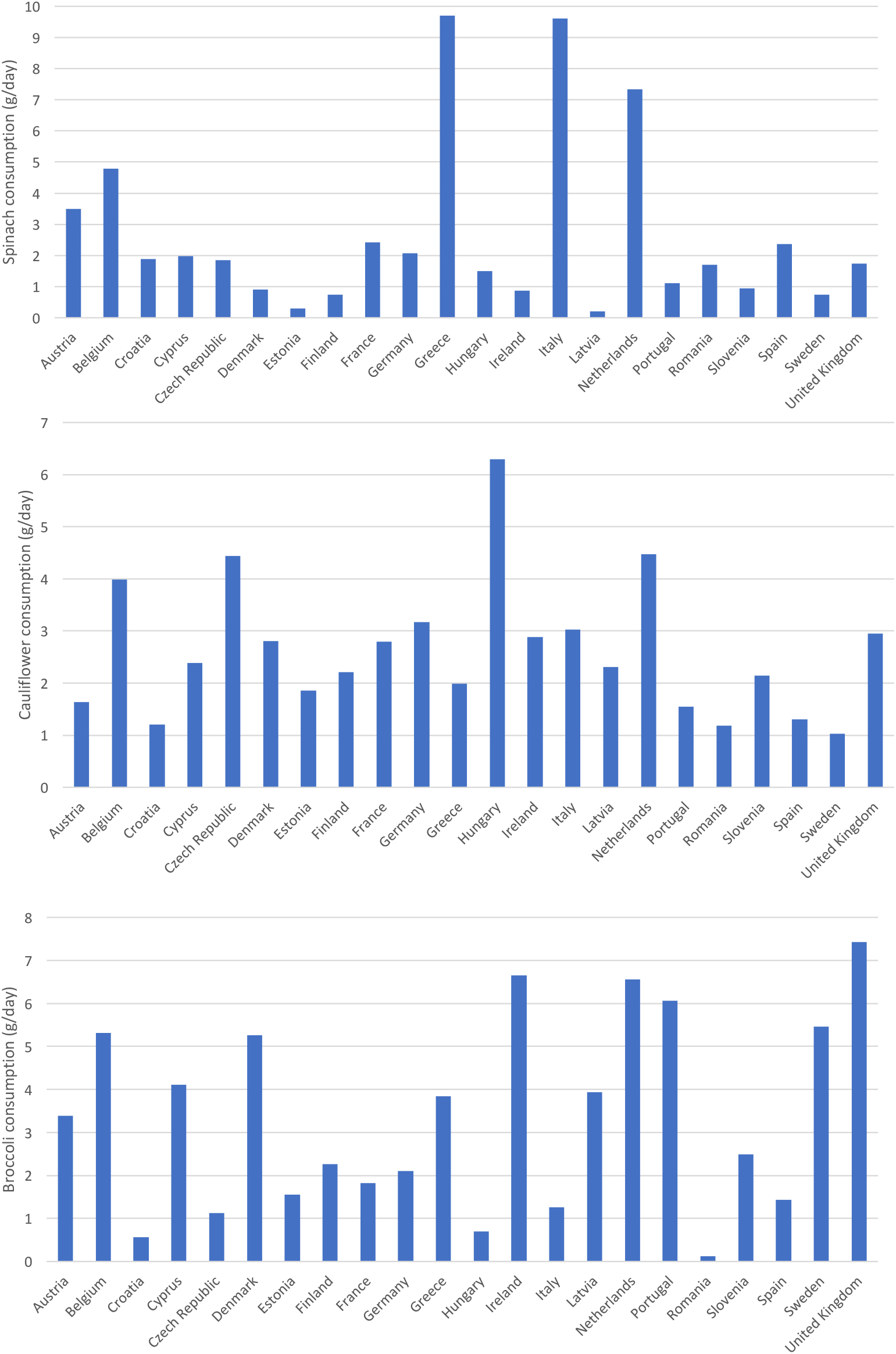
Consumption of different foods in Europe.

**Figure 2.**
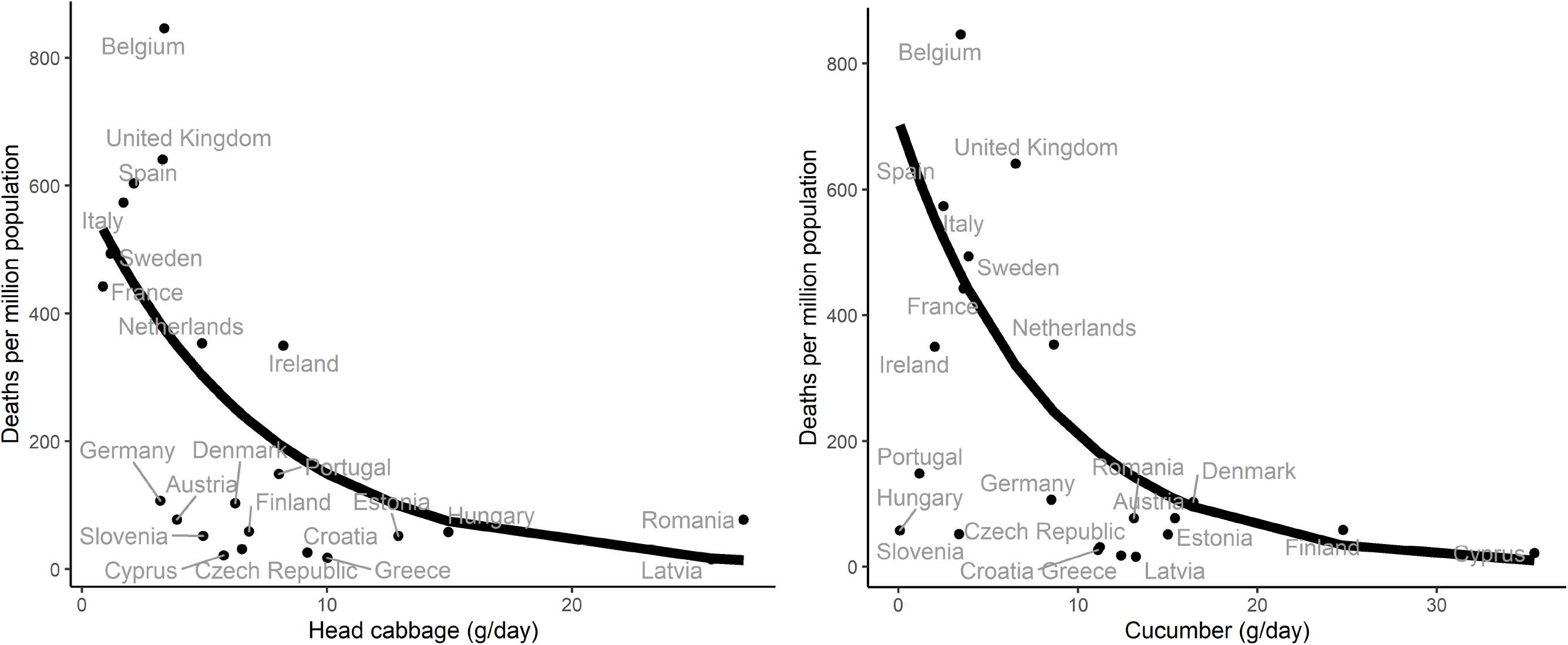
Raw data and estimated relationship between country death rates for COVID-19 and consumption of vegetables (g/day) using the quasi-Poisson regression model

After adjusting for potential confounders, for each g/day increase in consumption of head cabbage of the country, the mortality risk for COVID-19 decreases by 13.6 %. For each g/day increase in consumption of cucumber, the mortality risk decreases by 15.7%. Consumption of lettuce and broccoli showed the opposite pattern, i.e. a higher consumption was associated with a higher COVID-19 mortality. However, the results were more sensitive to adjustment, and the associations were no longer significant in some of the models.

## Discussion

This study shows that in countries where head cabbage and cucumber are highly consumed, death rates are low. On the other hand, there was no statistical significance on percentage change (p-value > 0.05) except for lettuce consumption and broccoli that were associated with an increased death rate only in some of the models.

### Limitations

According to the Johns Hopkins coronavirus resource center (https://coronavirus.jhu.edu), one of the most important ways of measuring the burden of COVID-19 is mortality. However, death rates are assessed differently between countries and there are many biases that are almost impossible to control ^1^. Using the rate of COVID-19 confirmed cases is subject to limitations. Differences in the mortality rates depend on the characteristics of the health care system, the reporting method, whether or not deaths outside the hospital have been counted and other factors, many of which remain unknown. Countries throughout the world have reported very different case fatality ratios - the number of deaths divided by the number of confirmed cases - but these numbers cannot be compared easily due to biases.

It is very important to consider differences in food consumption within countries but this cannot be studied using the EFSA database. As found in France, Spain and Italy, there are large regional differences in death rates and it would be of interest to compare sub-national regions with the different consumptions of fermented foods ^1^.

The consumption of vegetables is likely to show a seasonal pattern which may differ among countries. Our analysis used annual average consumption and a seasonal bias cannot be excluded.

A limited number of countries have been studied due to the lack of information on food consumption. A definite conclusion can therefore not be made. The selection of confounding factors has been arbitrary and more studies are needed. As in any ecological study, any inference from the observed association should be made at the country level, as the possibility of ecological fallacy precludes inferences at the individual level. Further testing in properly designed individual studies would be of interest.

Moreover, associations do not mean causality, and patterns of food consumption can be a proxy for other undetermined factors.

Understanding the multiple factors that are likely to explain inter and intra-national geographical variations will not be possible until indicators of the pandemic distribution have been improved and properly-designed studies conducted. However, this study is of interest since it is the first to attempt to link death rates with food consumption at a country level.

## Interpretation

COVID-19, like most diseases, exhibits large geographical variations which frequently remain unexplained despite abundant research ^8^. Though the more relevant factors are likely to be related to intensity and timing of interventions ^9^, demographic factors, seasonal variations, immunity, and cross-immunity, other factors like environment or nutrition should not be overlooked. Our study suggests that European countries with a lower COVID-19 mortality are more likely to exhibit a large consumption of head cabbage and cucumber.

Though our results do not allow to infer causality, they do reinforce our *a priory* hypothesis that the ingestion of anti-oxidant foods acting on insulin intolerance may have reduced the severity of COVID-19 and are in line with recent data on fermented foods ^6^. A large number of vegetables have an antioxidant activity and are effective in cardiovascular diseases ^10^ or diabetes ^11^. Antioxidants include mono- and polyunsaturated fatty acids, omega-3 fatty acids, antioxidant vitamins, minerals, phytochemicals, fiber, and plant protein ^12^.

Several foods can interact with transcription factors related to antioxidant effects such as Nrf2 ^13,14^. Natural compounds derived from vegetables such as curcumin, sulforaphane, resveratrol and vitamin D, among others, can activate Nrf2 ^15^. Cruciferous vegetables like Brassicaceae contain high amounts of sulforaphane, a potent activator of Nfr2 ^16^. However, broccoli ^17-19^ and cauliflower ^20^ have a potent anti-oxidant activity, but, in the present study, their consumption is not associated with differences in COVID-19 death rates between countries. It is possible that the amount ingested is too low to provide any protection. The other hypothesis that comes to mind here is that broccoli and cauliflower could be consumed cooked more often than raw ^21^. It should be noted that cucumber and head cabbage are reducing death rates for a consumption of over 10 g/day, whereas broccoli, leafy brassica and cauliflower are ingested at a dose of under 6 g/day. It is therefore possible that broccoli may have some effect at the individual level. Cucumber does not belong to the Brassicacea family but did however show a protective effect. Although less well studied, cucumber acts as an antioxidant on insulin resistance and its related diseases ^22-24^. Cucurbitacin B, a natural compound that exists in edible plants (bitter melons, cucumbers, pumpkins and zucchini), has anti-inflammatory and hypoglycemic properties through the activation of Nrf2 ^25,26^. These results and those of the recent study on fermented foods suggest a strong link between Nrf2 and the protection against severe forms of COVID-19.

The antioxidant effect of tomatoes or lettuce may be related to Nrf2, and, in the present study, may not be involved in the prevention of COVID-19 deaths. On the other hand, lettuce appears to increase death rates for unclear reasons.

Our study was restricted to European countries, so its results cannot be extrapolated to other regions. Thus, although this study is only indicative of the role of diet in COVID-19, it is however another piece of the hypothesis proposing that some vegetables with antioxidant properties may be involved in the prevention of severe COVID-19 at a country level.

## Data Availability

available under request

